# Association of Ventricular Arrhythmias with Lamotrigine: An Observational Cohort Study

**DOI:** 10.1101/2024.09.10.24313446

**Authors:** Sodam Kim, Landon Welch, Bertha De Los Santos, Przemysław B. Radwański, Mark A. Munger, Kibum Kim

## Abstract

**Background:** Whether lamotrigine (LTG) is associated with ventricular tachycardia (VT) in bipolar disorder (BPD), partial seizures (PSZ) and generalized tonic-clonic seizures (GTSZ) with and without structural heart disease (SHD) remains controversial. A mechanistic rational for LTG-induced re-entrant cardiac arrhythmias has recently been elucidated, leading to a real-world comparative cohort observational study being warranted.

**Methods:** A retrospective observational comparative safety study was performed using a large healthcare claims database of adult participants analyzing the one-year cumulative VT. Analytic cohort included adult participants diagnosed with bipolar I disorder (BPD), partial seizures (PSZ) or generalized tonic-clonic seizures (GTSZ). Participants were free from supraventricular or ventricular arrhythmias during the 6-month baseline period before the index LTG or CTR date. Exposure to LTG versus commonly prescribed alternative agents were the control comparators (CTR). One-year cumulative ventricular tachycardia (VT) incidence was calculated separately for GTSZ, PSZ and BPD using Kaplan-Meier estimator, with participants being censored at last enrollment, treatment switching or discontinuation. The VT association hazard ratios (HR) for LTG versus CTR was adjusted for baseline characteristics.

**Results:** The analytic cohort included 153,852 LTG and 213,593 CTR for BPD, 10,275 LTG and 24,971 CTR for PSZ, and 5,860 LTG and 17,506 CTR for GTSZ. Baseline cardiovascular risk profiles were higher among CTR than LTG across the three sub-analytic cohorts. The 1-year VT cumulative incidence from LTG or CTR free from was 0.79% vs 0.68% in BPD, 0.76% vs 0.58% in PSZ, and 0.93% vs 0.40% in GTSZ cohorts, The adjusted HR [95% CI] estimates were 1.326 [1.122-1.568, p<0.01], 1.403 [0.920-2.138, p=0.11], and 1.180 [0.607-2.295, p=0.63].

**Conclusions:** In adult participants, LTG has a strong association to increase VT risk compared to commonly prescribed alternatives.

**KEY POINTS:** **Question:** Does lamotrigine investigated in a real-world database increase the risk of ventricular tachycardia in patients with epilepsy or bipolar disease?

**Findings:** The lamotrigine-ventricular tachycardia association was statistically significant in adult bipolar disease participants. Although limited statistical significance, the positive association is ubiquitous across epileptic conditions. Structural heart disease has a notable increased effect on the incidence on the onset of ventricular tachycardia.

**Meaning:** Caution should be exercised in the use of lamotrigine in adult bipolar disease patients to avoid ventricular tachycardia.

## INTRODUCTION

Lamotrigine (LTG) is widely prescribed in the U.S., with thirteen million annual prescriptions for two million children, adolescents and adults.^1^ The drug has been established as first-line therapy for primary generalized tonic-clonic seizures and bipolar I disorder.^2–4^ The mechanism of action of LTG is not completely elucidated, but is believed to selectively bind and inhibit voltage-gated sodium channels (I_Na_), stabilizing presynaptic neuronal membranes and inhibiting presynaptic glutamate release.^5–6^ Case reports of lamotrigine-associated electrocardiographic events in epilepsy and bipolar disorder patients, with and without structural heart disease, have been published.^7–10^ Paired with recent publications that the inhibitory effect of LTG on I_Na_ blockade can occur at effective plasma concentrations that lead to pro-arrhythmia events,^11–12^ prompted the United States Food and Drug Association (FDA) to issue a warning that “*Lamictal exhibits class IB antiarrhythmic activity at therapeutically relevant concenttrations”*.^13^ This warning was issued due to evidence of LTG slowing conduction velocity, widening the QRS interval, leading to higher risk of pro-arrhythmic events in patients with structural heart disease and/or myocardial ischemia, but not in healthy subjects.^13^ New findings suggest that LTG may have enhanced affinity for the I_Na_ which may, in part, explain the potential for a clinical arrhythmogenic phenotype noted in epilepsy and possibly in bipolar disorders.^11,14^ However, not all data has shown a significance difference in arrhythmogenesis with lamotrigine.^14–15^ The current practice remains to be vigilant to the effects of LTG and consult a cardiologist before starting the agent, especially in patients with structural heart disease.^13^

To this end, a large population-based real-world study was designed to determine if LTG monotherapy was associated with ventricular tachycardia incidence in participants with and without structural heart disease treated for partial-, complex-seizures and bipolar disorders. Addressing this knowledge gap appears to be crucial to confirm the safety for millions of people prescribed LTG.

## METHODS

### Study design and Participants

This study is a retrospective comparative safety cohort study using Merative Health MarketScan® Commercial Claims and Medicare Supplemental databases (Merative L.P., Ann Arbor, MI). The data source covered medical and outpatient pharmacy claims from January 2009 to December 2021. Direct patient identifiers were removed before investigators had access to the data. The University of Illinois at Chicago Institutional Review Board deemed the use of the database for this study exempt.

Participants were defined based on the major LTG indications, namely bipolar I disorder (BPD), partial seizure (PSZ), or generalized tonic-clonic seizure (GTSZ), defined by the International classification of Diseases Ninth revision (ICD-9: 345.4, 345.5, 345.1, 296 and subcodes) or Tenth revision (ICD-10: G40.0, G40.1, G40.2, G40.6, F31 and subcodes) as a diagnosis of respective condition at either inpatient or outpatient setting. Eligible subjects must have had a record of dispensing LTG to be assigned to the LTG exposure cohort (LTG) or dispensing alternative medications indicated for respective condition to become a control subject (CTR): lithium, quetiapine, valproate or risperidone for BPD; carbamazepine, levetiracetam, oxcarbazepine or eslicarbazepine for PSZ; valproate, levetiracetam, carbamazepine or zonisamide for GTSZ. Prescription dispensing for each drug was defined by the National Drug Codes and generic name available from the outpatient pharmacy service records. The index date was defined by the first dispensing of the respective medication with a presence of BPD, PSZ, or GTSZ diagnosis within 180-day baseline period, without any record of exposure or comparator during the baseline period. The index medication had to be given at least seven days or longer without crossover between LTG and CTR.

To minimize confounding by pre-existing condition on the measure of association, we excluded participants who had ventricular tachycardia (VT), during the baseline period as defined by diagnosis codes (ICD-9 427 and 780.2 including all subcodes; ICD-10 I47, I48, I49, and R55 including all subcodes). Children or adolescents (< 18 years) at the index date were excluded from the analytic cohort.

### Baseline Characteristics, Outcome and Follow-up

Participant characteristics were collected at index date and during the 180-day baseline period. Demographics include age, sex, geographic location, and type of insurance. Clinical characteristics included chronic conditions and past/concurrent medications related to an arrhythmia or conditions to which the FDA LTG warning applied. We used diagnosis codes for the Charlson Comorbidity Index score and other relevant conditions (i.e., hypertension, hypercholesterolemia, depression, anxiety, substance abuse, eating disorders), and generic medication names or medication categories (angiotensin-converting enzyme inhibitors [ACEi], angiotensin II receptor blockers [ARB], non-dehydropyridine calcium channel blockers [NDHP-CCB], ßeta-adrenergic blockers [BB], spironolactone [SPL], and statins).^16–18^ The impact of structural heart disease (SHD) on the association between LTG and VT^13^ were applied (Supplement material, Table S1).

The outcome of interest was the onset of VT following the exposure to LTG or CTR for 7 days or longer period. The outcome was defined by ICD-9 (427.4x, 427.8x, 427.9x and 780.2x) or ICD-10 CM (I49.x and R55.x) diagnosis code for VT in any position from an outpatient service encounter, or in the first four positions from inpatient admission encounter. We followed the healthcare records for up to 365 days from the index date to assess the outcome. All participants met at least 7-day enrollment from the index date, during which subjects were free from the outcome of interest or the end of enrollment. Participants were required to have ≥ 7 days of medication supply.

### Statistical Analysis

Descriptive statistics and bivariate analyses were conducted to compare baseline characteristics between LTG and CTR cohorts. Summary statistics of age in year were mean and standard deviation (SD), which were compared using student t-test. Age was grouped into four categories of 18-25, 25-44, 45-64, and ≥ 65 years. All the other categorical variables were summarized using frequency (n) and percentage (%) out of the total number of participants in each cohort. We used Chi-square test for the statistical comparison of the categorical variables.

Time to VT and cumulative incidence was analyzed using a Kaplan-Meier estimator, for which participants were censored with treatment switching between LTG and CTR medications, the last date with available medication as defined by the days of supply from the most recent dispensing, or end of one-year follow up, whichever incurred first. CTR patient follow-up continued irrespective of drug switching among the control medications.

The measure of association was hazard ratio (HR). We calculated the HR and 95% confidence interval [95CI] of VT for LTG versus CTR using Cox-proportional hazard regression model. To adjust for the potential confounders, the multivariable regression included baseline characteristics that were significantly different (p<0.05) as covariates.

We assessed the impact of SHD on the risk of VT. First, we included SHD as part of covariate of the multivariable regression analysis with and without all the other covariates. The regression was followed by the inclusion of interaction between SHD and LTG, which allowed us to assess if the impact of LTG on the onset of VT differs by baseline SHD.

### Sensitivity Analysis – Propensity Score Matching

We assessed the sensitivity estimates and statistical conclusion to the analytic cohort definition for each of the three indications (BPD, PSZ, and GTSZ) using propensity score (PS) matching. The PS to receive LTG versus comparator were calculated using a logistic regression where all baseline characteristics were assessed as regressor on the stepwise selection process with p<0.15 as a selection criterion. LTG and CTR participants were exactly matched based on age group, sex, and propensity score caliper of 0.2 times of the standard deviation range. Cumulative incidence estimation using Kaplan-Meier approach and HR [95CI] calculation was undertaken using the PS-matched group where residual confounder after matching (standardized mean difference > 0.1) were adjusted.

## RESULTS

### Patient Characteristics

The three analytic cohorts for BPD, PSZ and GTSZ were made up of 367,445 (153,852 LTG and 213,593 CTR), 35,246 (10,275 LTG and 24,971 CTR), and 23,366 (5,860 LTG and 17,506 CTR) participants satisfying all inclusion and exclusion criteria, respectively (Figure 1). Across the BPD, PSZ and GTSZ cohorts, LTG participants were younger than CTR participants with the respective mean year age (SD) of 37.5 (13.6) vs. 40.9 (15.7), 41.3 (16.1) vs. 46.3 (17.5) and 36.8 (15.6) vs. 42.5 (17.6), and slightly more female dominated (69.7% vs. 59.9% BPD, 60.6% vs. 56.2% PSZ, and 60.0% vs. 53.2% GTSZ).

**FIGURE 1.**
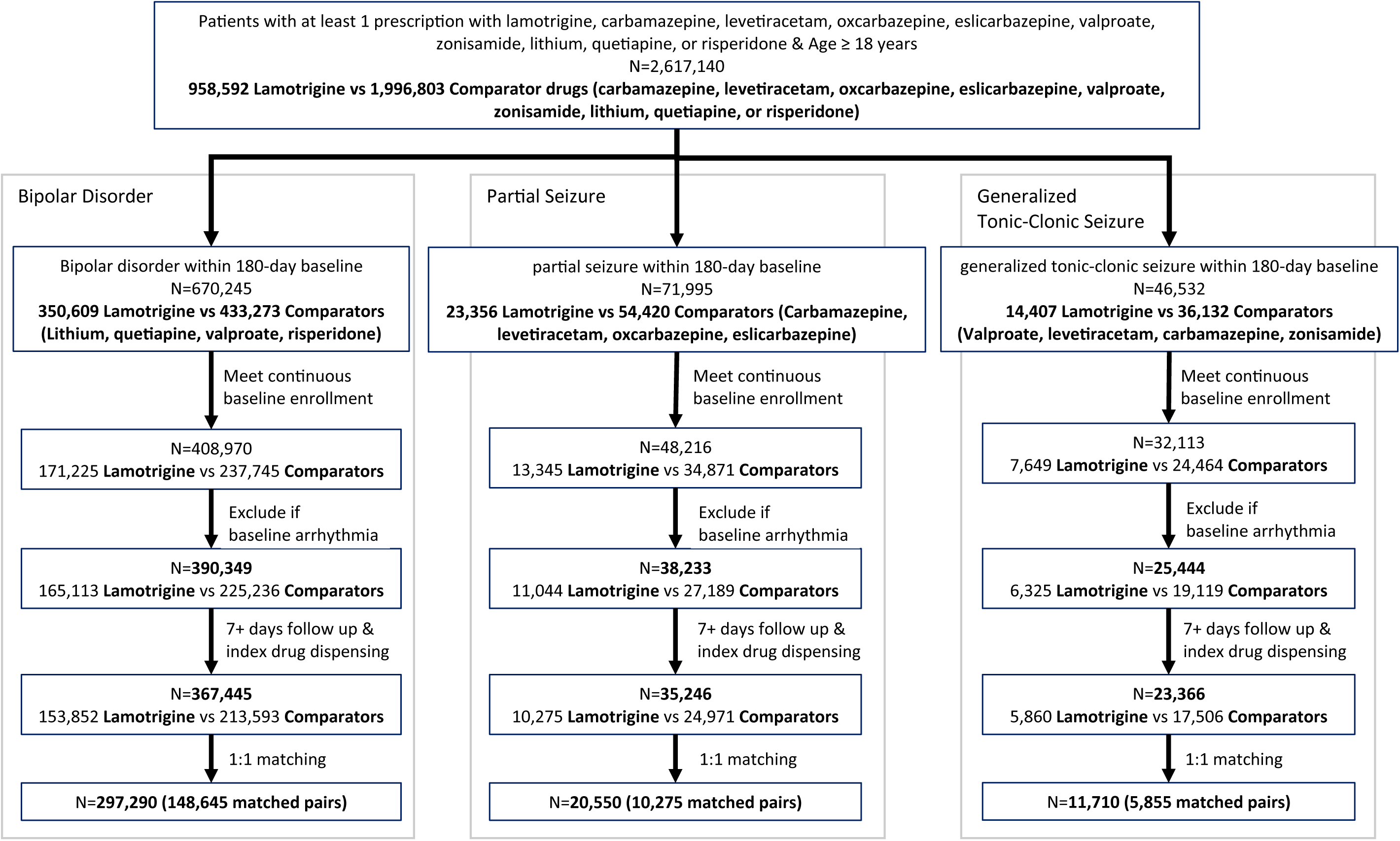
Patient Selection flow

**FIGURE 2.**
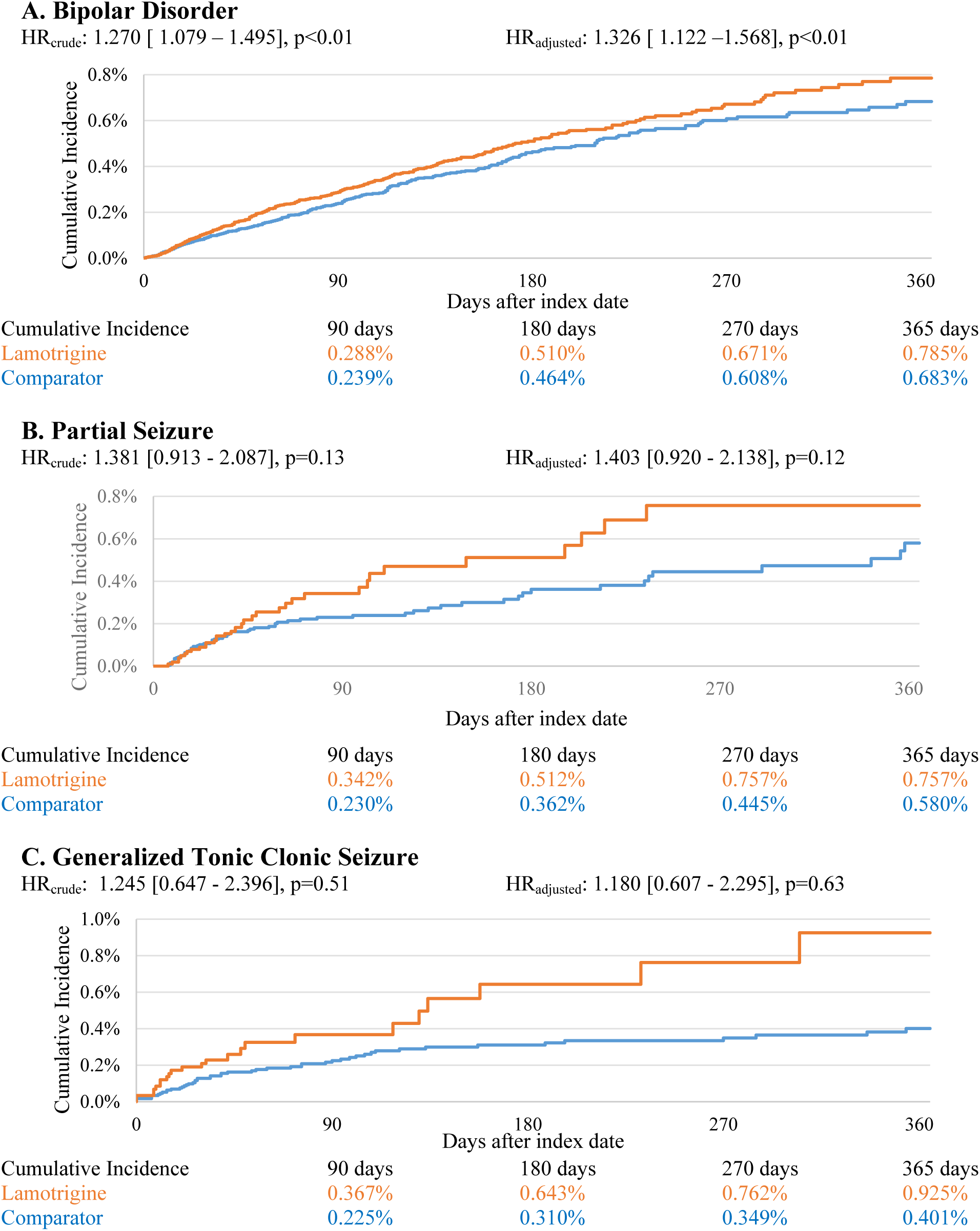
Kaplan-Meier Estimates of Cumulative Incidence of Vtach, Unmatched Analysis

**FIGURE 3.**
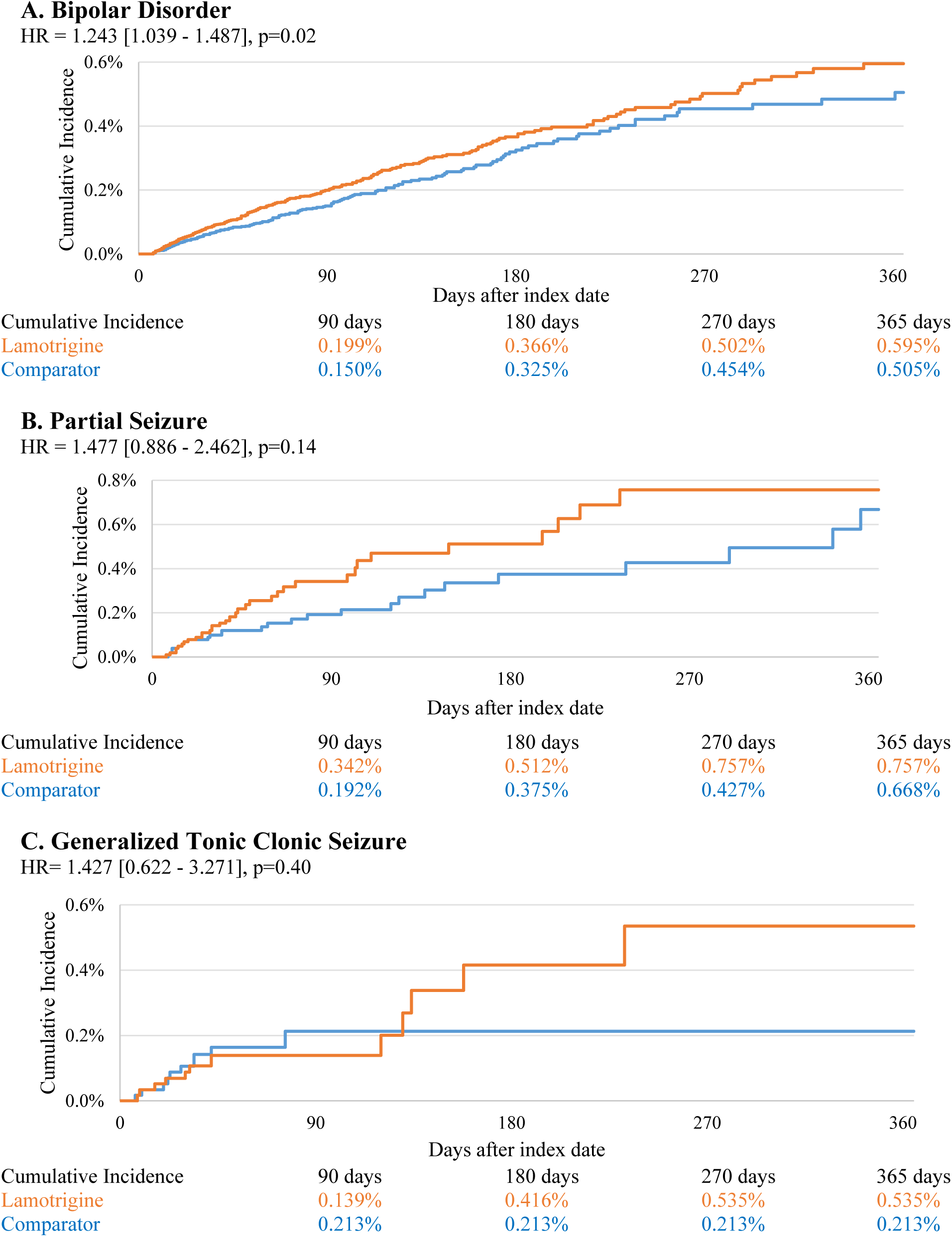
Kaplan-Meier Estimates of Cumulative Incidence of Vtach –Matched Analysis

While comorbidity profiles show that participants were generally healthy, the LTG cohort carried a lower risk of developing cardiovascular complications compared to the CTR cohort. For example, proportion of participants with history of cardiovascular disorder, including acute myocardial infarction (AMI), congestive heart failure (CHF), peripheral vascular disease, hypertension and hypercholesterolemia were lower in the LTG cohort versus CTR cohort with a strong statistical significance (p<0.01) across the all the three diagnostic groups.

Correspondingly, LTG participants were less likely to be prescribed ACEi, ARB, BB, NDHP-CCB and statin than CTR participants. Specifically, the prevalence of the composite SHD ^21^was significantly lower among LTG cohorts than CTR cohorts with respective prevalence’s of 2.2% vs. 4.2%, 6.9% vs. 13.5%, and 6.0% vs. 11.4% for BPD, PSZ and GTSZ. Anxiety and depression were more prevalent in the LTG than CTR among PSZ and GTSZ cohorts but were less prevalent in LTG than CTR among BPD cohort. (Table 1)

**TABLE 1.**
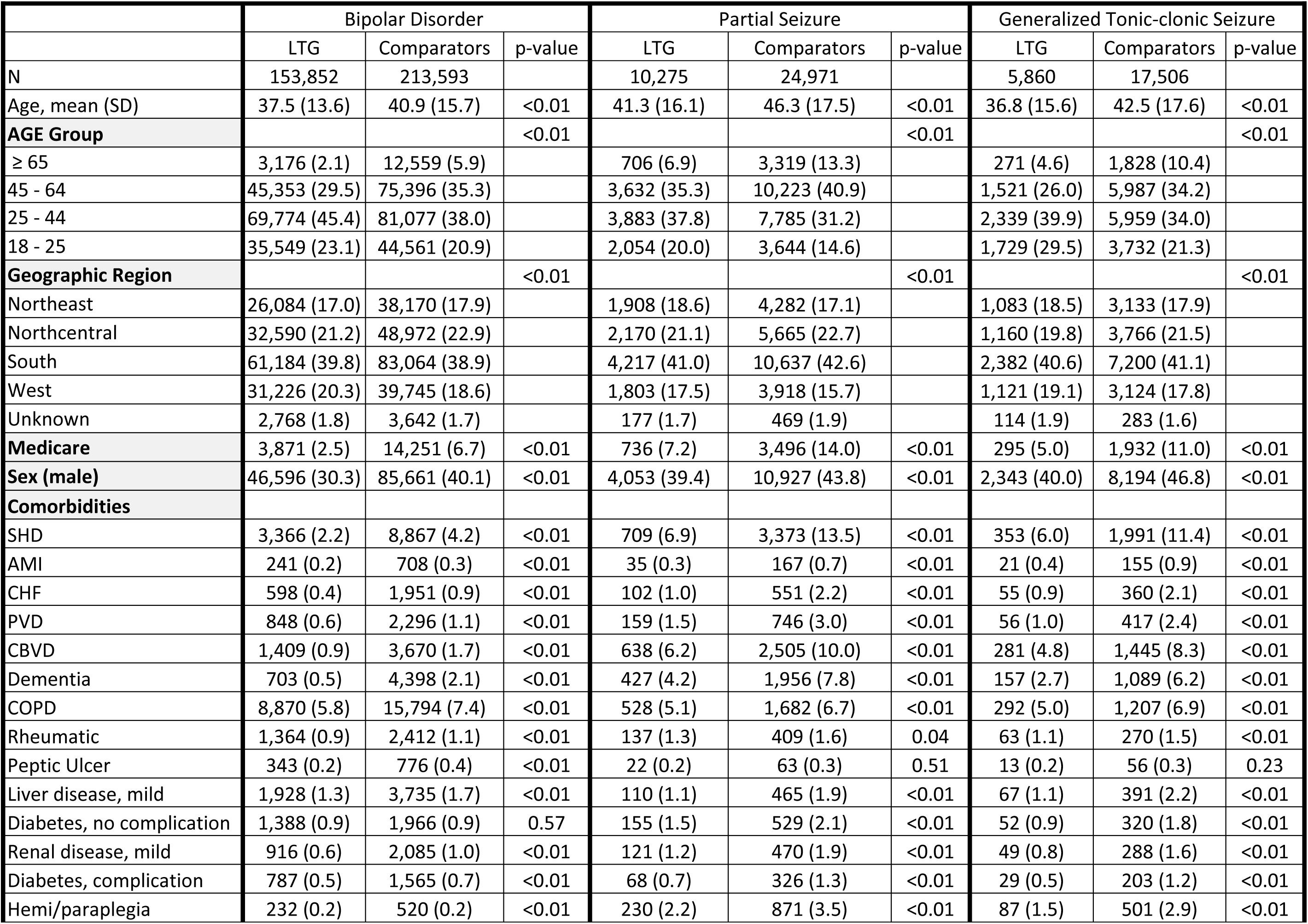
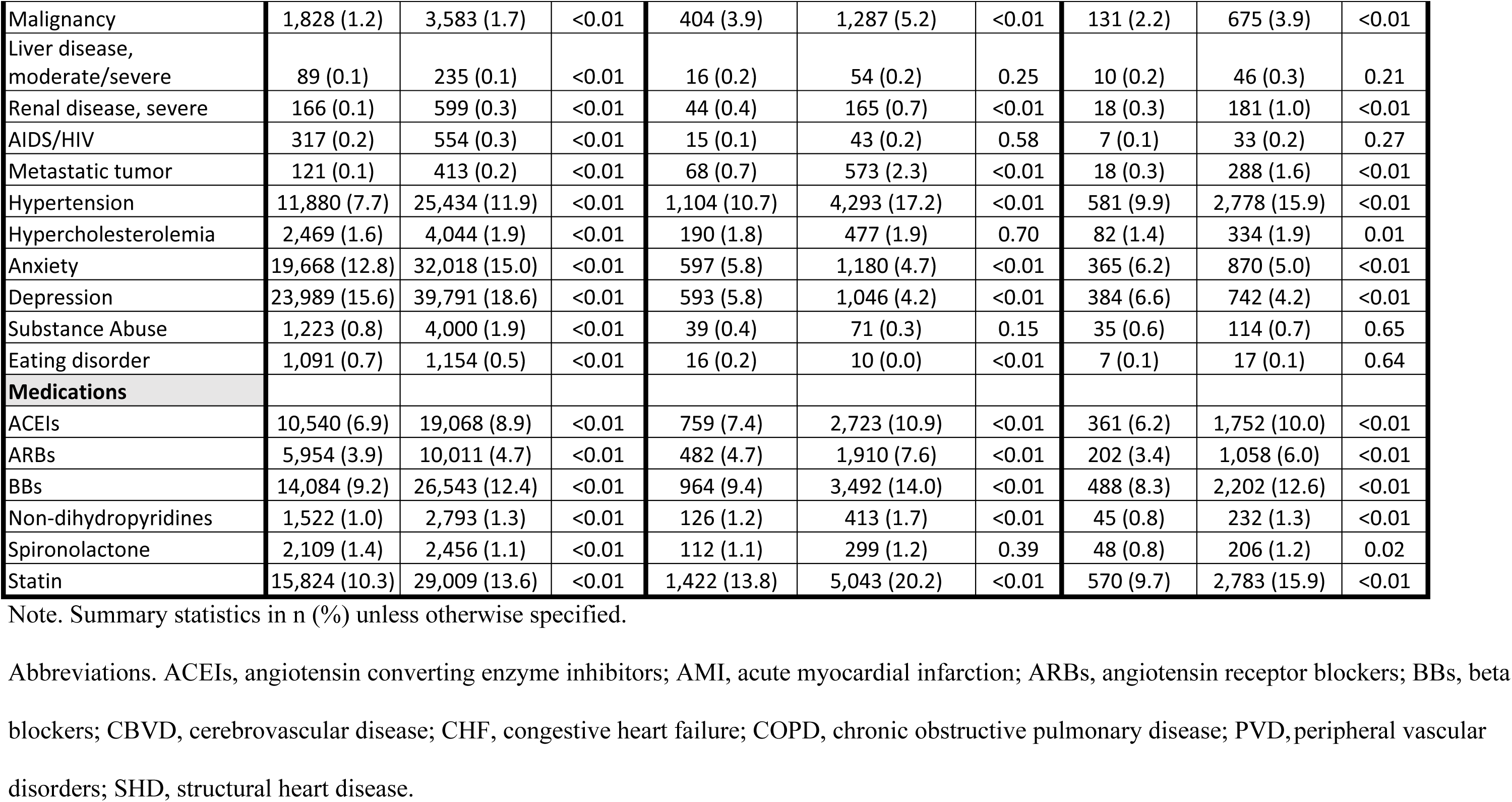
Baseline Characteristics

|  | Bipolar Disorder |  |  | Partial Seizure |  |  | Generalized Tonic-clonic Seizure |  |  |
| --- | --- | --- | --- | --- | --- | --- | --- | --- | --- |
|  | LTG | Comparators | p-value | LTG | Comparators | p-value | LTG | Comparators | p-value |
| N | 153,852 | 213,593 |  | 10,275 | 24,971 |  | 5,860 | 17,506 |  |
| Age, mean (SD) | 37.5 (13.6) | 40.9 (15.7) | <0.01 | 41.3 (16.1) | 46.3 (17.5) | <0.01 | 36.8 (15.6) | 42.5 (17.6) | <0.01 |
| **AGE Group** |  |  | <0.01 |  |  | <0.01 |  |  | <0.01 |
| ≥ 65 | 3,176 (2.1) | 12,559 (5.9) |  | 706 (6.9) | 3,319 (13.3) |  | 271 (4.6) | 1,828 (10.4) |  |
| 45 - 64 | 45,353 (29.5) | 75,396 (35.3) |  | 3,632 (35.3) | 10,223 (40.9) |  | 1,521 (26.0) | 5,987 (34.2) |  |
| 25 - 44 | 69,774 (45.4) | 81,077 (38.0) |  | 3,883 (37.8) | 7,785 (31.2) |  | 2,339 (39.9) | 5,959 (34.0) |  |
| 18 - 25 | 35,549 (23.1) | 44,561 (20.9) |  | 2,054 (20.0) | 3,644 (14.6) |  | 1,729 (29.5) | 3,732 (21.3) |  |
| **Geographic Region** |  |  | <0.01 |  |  | <0.01 |  |  | <0.01 |
| Northeast | 26,084 (17.0) | 38,170 (17.9) |  | 1,908 (18.6) | 4,282 (17.1) |  | 1,083 (18.5) | 3,133 (17.9) |  |
| Northcentral | 32,590 (21.2) | 48,972 (22.9) |  | 2,170 (21.1) | 5,665 (22.7) |  | 1,160 (19.8) | 3,766 (21.5) |  |
| South | 61,184 (39.8) | 83,064 (38.9) |  | 4,217 (41.0) | 10,637 (42.6) |  | 2,382 (40.6) | 7,200 (41.1) |  |
| West | 31,226 (20.3) | 39,745 (18.6) |  | 1,803 (17.5) | 3,918 (15.7) |  | 1,121 (19.1) | 3,124 (17.8) |  |
| Unknown | 2,768 (1.8) | 3,642 (1.7) |  | 177 (1.7) | 469 (1.9) |  | 114 (1.9) | 283 (1.6) |  |
| **Medicare** | 3,871 (2.5) | 14,251 (6.7) | <0.01 | 736 (7.2) | 3,496 (14.0) | <0.01 | 295 (5.0) | 1,932 (11.0) | <0.01 |
| **Sex (male)** | 46,596 (30.3) | 85,661 (40.1) | <0.01 | 4,053 (39.4) | 10,927 (43.8) | <0.01 | 2,343 (40.0) | 8,194 (46.8) | <0.01 |
| **Comorbidities** |  |  |  |  |  |  |  |  |  |
| SHD | 3,366 (2.2) | 8,867 (4.2) | <0.01 | 709 (6.9) | 3,373 (13.5) | <0.01 | 353 (6.0) | 1,991 (11.4) | <0.01 |
| AMI | 241 (0.2) | 708 (0.3) | <0.01 | 35 (0.3) | 167 (0.7) | <0.01 | 21 (0.4) | 155 (0.9) | <0.01 |
| CHF | 598 (0.4) | 1,951 (0.9) | <0.01 | 102 (1.0) | 551 (2.2) | <0.01 | 55 (0.9) | 360 (2.1) | <0.01 |
| PVD | 848 (0.6) | 2,296 (1.1) | <0.01 | 159 (1.5) | 746 (3.0) | <0.01 | 56 (1.0) | 417 (2.4) | <0.01 |
| CBVD | 1,409 (0.9) | 3,670 (1.7) | <0.01 | 638 (6.2) | 2,505 (10.0) | <0.01 | 281 (4.8) | 1,445 (8.3) | <0.01 |
| Dementia | 703 (0.5) | 4,398 (2.1) | <0.01 | 427 (4.2) | 1,956 (7.8) | <0.01 | 157 (2.7) | 1,089 (6.2) | <0.01 |
| COPD | 8,870 (5.8) | 15,794 (7.4) | <0.01 | 528 (5.1) | 1,682 (6.7) | <0.01 | 292 (5.0) | 1,207 (6.9) | <0.01 |
| Rheumatic | 1,364 (0.9) | 2,412 (1.1) | <0.01 | 137 (1.3) | 409 (1.6) | 0.04 | 63 (1.1) | 270 (1.5) | <0.01 |
| Peptic Ulcer | 343 (0.2) | 776 (0.4) | <0.01 | 22 (0.2) | 63 (0.3) | 0.51 | 13 (0.2) | 56 (0.3) | 0.23 |
| Liver disease, mild | 1,928 (1.3) | 3,735 (1.7) | <0.01 | 110 (1.1) | 465 (1.9) | <0.01 | 67 (1.1) | 391 (2.2) | <0.01 |
| Diabetes, no complication | 1,388 (0.9) | 1,966 (0.9) | 0.57 | 155 (1.5) | 529 (2.1) | <0.01 | 52 (0.9) | 320 (1.8) | <0.01 |
| Renal disease, mild | 916 (0.6) | 2,085 (1.0) | <0.01 | 121 (1.2) | 470 (1.9) | <0.01 | 49 (0.8) | 288 (1.6) | <0.01 |
| Diabetes, complication | 787 (0.5) | 1,565 (0.7) | <0.01 | 68 (0.7) | 326 (1.3) | <0.01 | 29 (0.5) | 203 (1.2) | <0.01 |
| Hemi/paraplegia | 232 (0.2) | 520 (0.2) | <0.01 | 230 (2.2) | 871 (3.5) | <0.01 | 87 (1.5) | 501 (2.9) | <0.01 |
| Malignancy | 1,828 (1.2) | 3,583 (1.7) | <0.01 | 404 (3.9) | 1,287 (5.2) | <0.01 | 131 (2.2) | 675 (3.9) | <0.01 |
| Liver disease, moderate/severe | 89 (0.1) | 235 (0.1) | <0.01 | 16 (0.2) | 54 (0.2) | 0.25 | 10 (0.2) | 46 (0.3) | 0.21 |
| Renal disease, severe | 166 (0.1) | 599 (0.3) | <0.01 | 44 (0.4) | 165 (0.7) | <0.01 | 18 (0.3) | 181 (1.0) | <0.01 |
| AIDS/HIV | 317 (0.2) | 554 (0.3) | <0.01 | 15 (0.1) | 43 (0.2) | 0.58 | 7 (0.1) | 33 (0.2) | 0.27 |
| Metastatic tumor | 121 (0.1) | 413 (0.2) | <0.01 | 68 (0.7) | 573 (2.3) | <0.01 | 18 (0.3) | 288 (1.6) | <0.01 |
| Hypertension | 11,880 (7.7) | 25,434 (11.9) | <0.01 | 1,104 (10.7) | 4,293 (17.2) | <0.01 | 581 (9.9) | 2,778 (15.9) | <0.01 |
| Hypercholesterolemia | 2,469 (1.6) | 4,044 (1.9) | <0.01 | 190 (1.8) | 477 (1.9) | 0.70 | 82 (1.4) | 334 (1.9) | 0.01 |
| Anxiety | 19,668 (12.8) | 32,018 (15.0) | <0.01 | 597 (5.8) | 1,180 (4.7) | <0.01 | 365 (6.2) | 870 (5.0) | <0.01 |
| Depression | 23,989 (15.6) | 39,791 (18.6) | <0.01 | 593 (5.8) | 1,046 (4.2) | <0.01 | 384 (6.6) | 742 (4.2) | <0.01 |
| Substance Abuse | 1,223 (0.8) | 4,000 (1.9) | <0.01 | 39 (0.4) | 71 (0.3) | 0.15 | 35 (0.6) | 114 (0.7) | 0.65 |
| Eating disorder | 1,091 (0.7) | 1,154 (0.5) | <0.01 | 16 (0.2) | 10 (0.0) | <0.01 | 7 (0.1) | 17 (0.1) | 0.64 |
| **Medications** |  |  |  |  |  |  |  |  |  |
| ACEIs | 10,540 (6.9) | 19,068 (8.9) | <0.01 | 759 (7.4) | 2,723 (10.9) | <0.01 | 361 (6.2) | 1,752 (10.0) | <0.01 |
| ARBs | 5,954 (3.9) | 10,011 (4.7) | <0.01 | 482 (4.7) | 1,910 (7.6) | <0.01 | 202 (3.4) | 1,058 (6.0) | <0.01 |
| BBs | 14,084 (9.2) | 26,543 (12.4) | <0.01 | 964 (9.4) | 3,492 (14.0) | <0.01 | 488 (8.3) | 2,202 (12.6) | <0.01 |
| Non-dihydropyridines | 1,522 (1.0) | 2,793 (1.3) | <0.01 | 126 (1.2) | 413 (1.7) | <0.01 | 45 (0.8) | 232 (1.3) | <0.01 |
| Spironolactone | 2,109 (1.4) | 2,456 (1.1) | <0.01 | 112 (1.1) | 299 (1.2) | 0.39 | 48 (0.8) | 206 (1.2) | 0.02 |
| Statin | 15,824 (10.3) | 29,009 (13.6) | <0.01 | 1,422 (13.8) | 5,043 (20.2) | <0.01 | 570 (9.7) | 2,783 (15.9) | <0.01 |

Note. Summary statistics in n (%) unless otherwise specified.
Abbreviations. ACEIs, angiotensin converting enzyme inhibitors; AMI, acute myocardial infarction; ARBs, angiotensin receptor blockers; BBs, beta blockers; CBVD, cerebrovascular disease; CHF, congestive heart failure; COPD, chronic obstructive pulmonary disease; PVD, peripheral vascular disorders; SHD, structural heart disease.

### Outcome

Cumulative incidence of VT at 12 months among LTG was 0.79%. 0.76% and 0.93% in BPD, PSZ, and GTSZ cohorts, respectively, which were higher than the estimates from the CTR cohort with the respective results (p-value from log-rank test) of 0.68% (0.0096), 0.58% (0.1245), and 0.40% (0.0092). (Figure 1) The difference in the cumulative incidence resulted in the crude HR [95CI] of VT for LTG versus CTR of 1.270 [1.079 - 1.495], 1.381 [0.913 - 2.087], and 1.245 [0.647 - 2.396], respectively. After statistical adjustment using a multivariable regression model, the results still indicated an increased risk of VT among LTG in BPD, but not in PSZ or GTSZ, with the respective HR [95CI] estimates for BPD, PSZ, and GTSZ of 1.326 [1.122 - 1.568], 1.403 [0.920 - 2.138], and 1.180 [0.607 - 2.295]. (Figure 1)

Our analysis also demonstrated an increase in the risk of VT with SHD. When SHD was a single covariate of the LTG vs. CTR – VT association, the HR [95CI] for SHD for BPD, PSZ and GTSZ cohorts were 1.878 [1.339 - 2.635], 1.393 [0.802 - 2.417], 1.931 [0.937 - 3.981], respectively. SHD was positively associated with the onset of VT, but the 95CI started to cross the null when SHD was a part of the full multivariable regression model with the respective HR [95CI] of 1.538 [1.011 - 2.341], 1.539 [0.752 - 3.149], 2.056 [0.775 - 5.45]. Analysis including the interaction term resulted in a strong positive but statistically insignificant difference of SHD impact on LTG versus CTR (HR [95CI] of 1.615 [0.819 - 3.183], 1.35 [0.396 - 4.606], and 1.789 [0.319 - 10.031], respectively. (Table 2)

**TABLE 2.** Estimates for LTG, SHD and SHD – LTG interaction effects on the onset of VT.

| Regressor | Hazard ratio [95CI] from each regression coefficient |  |  |
| --- | --- | --- | --- |
|  | Bipolar Disorder | Partial Seizure | Generalized Tonic Clonic Seizure |
| LTG<sup>1</sup> | 1.270 [1.079 - 1.495] | 1.381 [0.913 - 2.087] | 1.245 [0.647 - 2.396] |
| LTG<sup>2</sup> | 1.296 [1.100 - 1.526] | 1.413 [0.933 - 2.141] | 1.303 [0.675 - 2.513] |
| SDH<sup>2</sup> | 1.878 [1.339 - 2.635] | 1.393 [0.802 - 2.417] | 1.931 [0.937 - 3.981] |
| LTG<sup>3</sup> | 1.259 [1.064 - 1.490] | 1.362 [0.873 - 2.123] | 1.196 [0.584 - 2.449] |
| SDH<sup>3</sup> | 1.553 [0.986 - 2.446] | 1.290 [0.676 - 2.462] | 1.727 [0.762 - 3.913] |
| LTG-SDH interaction<sup>3</sup> | 1.615 [0.819 - 3.183] | 1.350 [0.396 - 4.606] | 1.789 [0.319 - 10.031] |
| Model: 1 $h(t)=h_0(t) \exp (\beta_{LTG} \cdot x_{LTG})$; 2. $h(t)=h_0(t) \exp (\beta_{LTG} \cdot x_{LTG} + \beta_{SHD} \cdot x_{SHD})$; 3. $h(t)=h_0(t) \exp (\beta_{LTG} \cdot x_{LTG} + \beta_{SHD} \cdot x_{SHD} + \beta_{LTG*SHD} \cdot x_{LTG*SHD})$ | | | |

### Sensitivity Analysis

A matched cohort was created with 148,645, 10,275 and 5,855 pairs for BPD, PSZ and GTSZ, respectively. The propensity score and exact matching criteria relieved concerns on the confounding by patient characteristics with absolute standardized mean differences is equal to or less than 0.1 for all the variables except North-East region and baseline metastatic solid for the PSZ cohort (Supplement material, Table S2).

The rate of the onset of VT was higher among those exposed to LTG compared to comparators, with 365-day cumulative incidence estimates of 0.60 vs. 0.51% (log-rank test, p=0.0173) in the BPD, 0.76 vs. 0.67% (log-rank test, p=0.1323) in the PSZ, and 0.54 vs. 0.21% (log-rank test, p=0.3987) in the GTSZ cohort. The respective HR [95CI] for LTG versus CTR at 12-month follow-up were applied the HR [95CI] were 1.243 [1.039 - 1.487], 1.477 [0.886 - 2.462], and 1.427 [0.622 – 3.271], respectively.

## DISCUSSION

In a large observational cohort study from a United States-based commercial insurance and Medicare supplement claims database, LTG exposure was associated with an increase in the cumulative incidence of ventricular tachycardia across the diagnostic categories of partial-, complex-seizures, and bipolar disorders when compared to commonly used agents. After adjustment for multivariate regression model, LTG use in BPD remained significant, but not for the epilepsy cohorts. The association was significant when controlled for SHD as a single covariate but was insignificant when the multivariate model included SHD on top of the other covariates. The increase in ventricular tachycardia incidence was approximately 1% of the population. Most of the encounters with the diagnosis of VT occurred in the first 30 days.

This study is the first to report a higher risk of LTG-associated ventricular tachycardia in BPD participants, with and importantly without structural heart disease. These findings project the number needed to harm of 980, placing 3% of the 7 million bipolar disorder I persons ≥ 45 years, approximately 210,000 participants, at risk of LTG-associated ventricular tachycardia yearly.^19^ LTG-associated ventricular tachycardia may be more prevalent in males between 45 to 64 years compared to younger participants, findings previously noted.^9^ VT onset of approximately 20-30+ days (Figure 1), may be associated with up-titration of LTG dosage and administration guidelines in pre-disposed at-risk participants.^20^ Thereby, higher LTG concentrations with greater affinity for the I_Na,_ may contribute to enhanced arrhythmogenic phenotype in BPD patients.^11, 21–22^ Our finding of an increase in LTG-associated VT in BPD adult participants without structural heart disease maybe have been masked over time by high number of suicides recorded with BPD. BPD has the highest rate of suicide of all psychiatric conditions and is approximately 20–30 times that of the general population.^23^

LTG in participants without structural heart disease is a new finding given that healthy people undergoing QT testing have not shown changes in QRS duration,^24^ but QRS prolongation should be considered a risk for LTG ventricular tachycardia.^25^ Toxic concentrations of LTG (i.e., 7-10 fold higher than therapeutic concentrations) have been associated with QRS prolongation and/or development of sustained ventricular tachycardia.^12^ LTG exhibits voltage-gated sodium channel use-dependent block properities^26^ supporting that rapid heart rates may be associated with ventricular tachycardia risk at human physiologic concentrations.^12^ QRS prolongation is partly determined by resistance across intracellular connections, or gap junctions, between cardiac myocytes.^27^ Ventricular gap junctions are constituted by connexin proteins with connexin isoform-43 being the predominant protein in humans.^28^ In studies of human cardiac gap junctions using a dye transfer “parachute assay” to determine IC_50_ values for compounds, LTG was found to uncouple connexin isoform-43.^29^ This activity is associated with proclivity for QRS prolongation and maybe dependent on direct gap junction excitation or by non-gap-junction-mediated ephaptic mechanisms.^30–31^ Thereby, LTG, a potent Na_v_1.5 blocker, may uncouple connexin 43, prolonging the QRS complex, causing ventricular tachycardia.

### Limitations

Interpreting our findings, readers should be mindful of several limitations. First, VT is correlated with mortality, ^32^ but we were unable to link mortality from the claim records to death certificates. Thereby, detected VT could be subject to the survivalship bias that needs to be determined and many patients who already have VT might not have a chance to claim the condition, which likely had a significant impact on the measure of association. Second, our study is subject to event size. To determine the statistical significance of the LTG versus CTR and its interaction with baseline characteristics, a sufficient number of events and absolute difference is required. Since our VT cumulative incidence estimates were less than 1% across the three diagnosis criteria, statistical interpretation is limited by the small number of events and effect size. Third, our study included a large number of regression variables, which raises concern on the potential interactions and collinearity among independent variables. Although this would not significantly change the magnitude or direction of the LTG-outcome association, future studies need to focus on the other risk factors that could modify the measure of association. Lastly, any interpretation should be considered considering the limitations of insurance claims. Our study findings and conclusion may be subject to miscoding and unobserved confounders. In addition, the study findings have limited generalizability to the commercially insured population. The validity of using diagnosis codes to determine VT event will be another theme of future research.

### Conclusions

This study provides a general insight into the positive real-world association between LTG use and VT. The LTG-VT association was clinically and statistically significant in adult BPD participants. Although limited statistical significance, the positive LTG-VT association is ubiquitous across epileptic conditions. SHD has a notable increased effect on the incidence on the onset of VT.

## Data Availability

Data is available via email to the authors

## Author Disclosures

The authors have no disclosures with the content of this manuscript.

## Author Contributions

All authors fully contributed to the content of this manuscript, including meeting the four criteria of the Internal Committee of Medical Journal Editors. All authors had full access to all the data in the study and took full responsibility for integrity of the work and accuracy of the data analysis, from inception to published article.

## Conflict of Interest

The authors have no conflict of interest associated with the content of this manuscript.

## Analytic Personnel

Sodam Kim, Pharm.D., Mark A. Munger, Pharm.D., Kibum Kim, Ph.D.

## Funding

National Institutes of Health from grants R01HL155378 and R01 NS121234 to Dr. Radwanski.

## NON-STANDARD ABBREVIATIONS AND ACRONYMS

ACEi: Angiotensin-converting enzyme inhibitor
ARB: Angiotensin receptor blocker
BB: ßeta-adrenergic blocker
BPD: Bipolar 1 Disorder
CTR: Control Group
FDA: U.S. Food and Drug Administration
GTSZ: Generalized tonic-clonic seizure
HR: Hazard Ratio
INa: Voltage-gated sodium channels
LTG: Lamotrigine
NDHP-CCB: Non-dihydropyridine Calcium Channel Blocker
PSZ: Partial Seizure
PS: Propensity Score
SHD: Structural Heart Disease
SPL: Spironolactone
SMD: Standard Mean Difference
QRS: QRS Complex
QT: QT Interval
VT: Ventricular Tachycardia

## ACKNOWLEDGEMENT

We express our gratitude to Marco Bortalato, M.D., Ph.D., Professor of Pharmacology and Toxicology, University of Utah for his expertise and counsel towards this manuscript.

## Supplement Material

**Table S1.** Diagnosis codes for Structured Heart Diseases.

|  | ICD-10 | ICD-9 |
| --- | --- | --- |
| Myocardial Ischemia | I21.A1 | 414.9 |
| SIHD | I24.9 | 414 |
| CAD | I25.10 | 414.01 |
| ACS Event | I20.0 | 411.1 |
| HF | I50.82 | 428 |
| Structural Heart Disease | Q24.9 | Not applicable |
| Cardiomyopathy | I42.9 | 425.4 |
| Heart valve disease | Aortic - I35.9 | Aortic - 395.0 |
|  | Tricuspid - I08.3 | Tricuspid - 397.0 |
|  | Mitral - I08.1 | Mitral - 394.0 |
|  | Any - I08.8 |  |
| Brugada syndrome | I49.8 | 426.81 |
| Sodium Channelopathies | E87.1 | Not applicable |
| Acute myocardial infarction | I21 | 410 |
| stroke | I63 | 434.91 |
| Hospitalization with HF | I50.9 | V66.7 |

**Supplement material - Table S2.** Baseline Characteristics after propensity score matching

|  | Bipolar Disorder |  |  |  | Partial Seizure |  |  |  | Generalized Tonic-clonic Seizure |  |  |  |
| --- | --- | --- | --- | --- | --- | --- | --- | --- | --- | --- | --- | --- |
|  | LTG | Comparators | p-value | SMD | LTG | Comparators | p-value | SMD | LTG | Comparators | p-value | SMD |
| N | 148,645 | 148,645 |  |  | 10,275 | 10,275 |  |  | 5,855 | 5,855 |  |  |
| Age, mean (SD) | 37.88 (13.65) | 38.30 (14.03) |  | -0.030 | 41.31 (16.12) | 41.77 (16.47) |  | -0.028 | 36.83 (15.56) | 37.44 (16.12) |  | -0.038 |
| **AGE Group** |  |  | 1.00 |  |  |  | 1.00 |  |  |  | 1.00 | 0.000 |
| ≥ 65 | 3,173 (2.1) | 3,173 (2.1) |  | 0.000 | 706 (6.9) | 706 (6.9) |  | 0.000 | 271 (4.6) | 271 (4.6) |  | 0.000 |
| 45 - 64 | 45,335 (30.5) | 45,335 (30.5) |  | 0.000 | 3,632 (35.3) | 3,632 (35.3) |  | 0.000 | 1,519 (25.9) | 1,519 (25.9) |  | 0.000 |
| 25 - 44 | 67,420 (45.4) | 67,420 (45.4) |  | 0.000 | 3,883 (37.8) | 3,883 (37.8) |  | 0.000 | 2,336 (39.9) | 2,336 (39.9) |  | 0.000 |
| 18 - 25 | 32,717 (22.0) | 32,717 (22.0) |  | 0.000 | 2,054 (20.0) | 2,054 (20.0) |  | 0.000 | 1,729 (29.5) | 1,729 (29.5) |  | 0.000 |
| **Geographic Region** |  |  | <0.01 |  |  |  | <0.01 |  |  |  | 1.00 | 0.000 |
| Northeast | 25,222 (17.0) | 24,866 (16.7) |  | 0.006 | 1,908 (18.6) | 1,496 (14.6) |  | 0.108 | 1,083 (18.5) | 1,082 (18.5) |  | 0.000 |
| Northcentral | 31,527 (21.2) | 34,159 (23.0) |  | -0.043 | 2,170 (21.1) | 2,370 (23.1) |  | -0.047 | 1,160 (19.8) | 1,160 (19.8) |  | 0.000 |
| South | 59,112 (39.8) | 59,639 (40.1) |  | -0.007 | 4,217 (41.0) | 4,561 (44.4) |  | -0.068 | 2,382 (40.7) | 2,382 (40.7) |  | 0.000 |
| West | 30,126 (20.3) | 28,073 (18.9) |  | 0.035 | 1,803 (17.5) | 1,702 (16.6) |  | 0.026 | 1,119 (19.1) | 1,121 (19.1) |  | -0.001 |
| Unknown | 2,658 (1.8) | 1,908 (1.3) |  | 0.041 | 177 (1.7) | 146 (1.4) |  | 0.024 | 111 (1.9) | 110 (1.9) |  | 0.001 |
| **Medicare** | 3,867 (2.6) | 4,437 (3.0) | <0.01 | -0.023 | 736 (7.2) | 789 (7.7) | 0.16 | -0.020 | 295 (5.0) | 302 (5.2) | 0.77 | -0.005 |
| **Sex (male)** | 46,580 (31.3) | 46,580 (31.3) | 1.00 | 0.000 | 4,053 (39.4) | 4,053 (39.4) | 1.00 | 0.000 | 2,343 (40.0) | 2,343 (40.0) | 1.00 | 0.000 |
| **Comorbidities** |  |  |  |  |  |  |  |  |  |  |  |  |
| SHD | 3,360 (2.3) | 3,393 (2.3) | 0.68 | -0.001 | 709 (6.9) | 978 (9.5) | <0.01 | -0.095 | 353 (6.0) | 445 (7.6) | <0.01 | -0.062 |
| AMI | 241 (0.2) | 262 (0.2) | 0.35 | -0.003 | 35 (0.3) | 44 (0.4) | 0.31 | -0.014 | 21 (0.4) | 34 (0.6) | 0.08 | -0.032 |
| CHF | 597 (0.4) | 713 (0.5) | <0.01 | -0.012 | 102 (1.0) | 143 (1.4) | <0.01 | -0.037 | 55 (0.9) | 63 (1.1) | 0.46 | -0.014 |
| PVD | 844 (0.6) | 954 (0.6) | <0.01 | -0.010 | 159 (1.5) | 213 (2.1) | <0.01 | -0.039 | 56 (1.0) | 98 (1.7) | <0.01 | -0.063 |
| CBVD | 1,407 (0.9) | 1,364 (0.9) | 0.41 | 0.003 | 638 (6.2) | 855 (8.3) | <0.01 | -0.081 | 280 (4.8) | 379 (6.5) | <0.01 | -0.073 |
| Dementia | 699 (0.5) | 1,628 (1.1) | <0.01 | -0.071 | 427 (4.2) | 550 (5.4) | <0.01 | -0.056 | 157 (2.7) | 239 (4.1) | <0.01 | -0.078 |
| COPD | 8,860 (6.0) | 8,910 (6.0) | 0.70 | -0.001 | 528 (5.1) | 622 (6.1) | <0.01 | -0.040 | 290 (5.0) | 341 (5.8) | 0.04 | -0.039 |
| Rheumatic | 1,364 (0.9) | 1,353 (0.9) | 0.83 | 0.001 | 137 (1.3) | 164 (1.6) | 0.12 | -0.022 | 62 (1.1) | 75 (1.3) | 0.26 | -0.021 |
| Peptic Ulcer | 342 (0.2) | 338 (0.2) | 0.88 | 0.001 | 22 (0.2) | 28 (0.3) | 0.40 | -0.012 | 13 (0.2) | 16 (0.3) | 0.58 | -0.010 |
| Liver disease, mild | 1,902 (1.3) | 2,341 (1.6) | <0.01 | -0.025 | 110 (1.1) | 173 (1.7) | <0.01 | -0.053 | 66 (1.1) | 97 (1.7) | 0.01 | -0.045 |
| Diabetes, no complication | 1,368 (0.9) | 1,226 (0.8) | <0.01 | 0.010 | 155 (1.5) | 150 (1.5) | 0.77 | 0.004 | 52 (0.9) | 65 (1.1) | 0.23 | -0.022 |
| Renal disease, mild | 908 (0.6) | 952 (0.6) | 0.31 | -0.004 | 121 (1.2) | 141 (1.4) | 0.21 | -0.017 | 49 (0.8) | 63 (1.1) | 0.18 | -0.025 |
| Diabetes, complication | 787 (0.5) | 727 (0.5) | 0.12 | 0.006 | 68 (0.7) | 80 (0.8) | 0.32 | -0.014 | 29 (0.5) | 60 (1.0) | <0.01 | -0.061 |
| Hemi/paraplegia | 224 (0.2) | 304 (0.2) | <0.01 | -0.013 | 230 (2.2) | 342 (3.3) | <0.01 | -0.066 | 87 (1.5) | 156 (2.7) | <0.01 | -0.083 |
| Malignancy | 1,813 (1.2) | 2,025 (1.4) | <0.01 | -0.013 | 404 (3.9) | 440 (4.3) | 0.21 | -0.018 | 131 (2.2) | 184 (3.1) | <0.01 | -0.056 |
| Liver disease, moderate/severe | 88 (0.1) | 123 (0.1) | 0.02 | -0.009 | 16 (0.2) | 26 (0.3) | 0.12 | -0.022 | 10 (0.2) | 5 (0.1) | 0.20 | 0.024 |
| Renal disease, severe | 166 (0.1) | 278 (0.2) | <0.01 | -0.020 | 44 (0.4) | 45 (0.4) | 0.92 | -0.001 | 18 (0.3) | 43 (0.7) | <0.01 | -0.059 |
| AIDS/HIV | 316 (0.2) | 341 (0.2) | 0.33 | -0.004 | 15 (0.1) | 19 (0.2) | 0.49 | -0.010 | 7 (0.1) | 11 (0.2) | 0.35 | -0.017 |
| Metastatic tumor | 121 (0.1) | 218 (0.1) | <0.01 | -0.019 | 68 (0.7) | 206 (2.0) | <0.01 | -0.117 | 18 (0.3) | 68 (1.2) | <0.01 | -0.100 |
| Hypertension | 11,783 (7.9) | 14,120 (9.5) | <0.01 | -0.056 | 1,104 (10.7) | 1,299 (12.6) | <0.01 | -0.059 | 580 (9.9) | 649 (11.1) | 0.04 | -0.038 |
| Hypercholesterolemia | 2,464 (1.7) | 2,435 (1.6) | 0.68 | 0.002 | 190 (1.8) | 161 (1.6) | 0.12 | 0.022 | 82 (1.4) | 74 (1.3) | 0.52 | 0.012 |
| Anxiety | 19,662 (13.2) | 19,686 (13.2) | 0.90 | 0.000 | 597 (5.8) | 518 (5.0) | 0.02 | 0.034 | 362 (6.2) | 359 (6.1) | 0.91 | 0.002 |
| Depression | 23,981 (16.1) | 24,026 (16.2) | 0.82 | -0.001 | 593 (5.8) | 412 (4.0) | <0.01 | 0.082 | 383 (6.5) | 264 (4.5) | <0.01 | 0.089 |
| Substance Abuse | 1,189 (0.8) | 2,619 (1.8) | <0.01 | -0.086 | 39 (0.4) | 31 (0.3) | 0.34 | 0.013 | 35 (0.6) | 48 (0.8) | 0.15 | -0.026 |
| Eating disorder | 953 (0.6) | 952 (0.6) | 0.98 | 0.000 | 16 (0.2) | 2 (0.0) | <0.01 | 0.046 | 7 (0.1) | 8 (0.1) | 0.80 | -0.005 |
| **Medications** |  |  |  |  |  |  |  |  |  |  |  |  |
| ACEIs | 10,448 (7.0) | 10,883 (7.3) | <0.01 | -0.011 | 759 (7.4) | 758 (7.4) | 0.98 | 0.000 | 361 (6.2) | 423 (7.2) | 0.02 | -0.042 |
| ARBs | 5,910 (4.0) | 5,476 (3.7) | <0.01 | 0.015 | 482 (4.7) | 568 (5.5) | <0.01 | -0.038 | 202 (3.5) | 272 (4.6) | <0.01 | -0.061 |
| BBs | 14,078 (9.5) | 14,098 (9.5) | 0.90 | 0.000 | 964 (9.4) | 965 (9.4) | 0.98 | 0.000 | 487 (8.3) | 563 (9.6) | 0.01 | -0.045 |
| Non-dihydropyridines | 1,522 (1.0) | 1,477 (1.0) | 0.41 | 0.003 | 126 (1.2) | 136 (1.3) | 0.53 | -0.009 | 43 (0.7) | 45 (0.8) | 0.83 | -0.004 |
| Spironolactone | 2,028 (1.4) | 1,744 (1.2) | <0.01 | 0.017 | 112 (1.1) | 119 (1.2) | 0.64 | -0.006 | 48 (0.8) | 54 (0.9) | 0.55 | -0.011 |
| Statin | 15,732 (10.6) | 15,740 (10.6) | 0.96 | 0.000 | 1,422 (13.8) | 1,422 (13.8) | 1.00 | 0.000 | 570 (9.7) | 597 (10.2) | 0.40 | -0.015 |

Note. Summary statistics in n (%) unless otherwise specified.
Abbreviations. ACEIs, angiotensin converting enzyme inhibitors; AMI, acute myocardial infarction; ARBs, angiotensin receptor blockers; BBs, beta blockers; CBVD, cerebrovascular disease; CHF, congestive heart failure; COPD, chronic obstructive pulmonary disease; PVD, peripheral vascular disorders; SHD, structural heart disease; SMD, standardized mean difference (valance statistics for the matched cohort)

## Notes

### Competing Interest Statement

The authors have declared no competing interest.

### Clinical Trial

This study is not a clinical trial.

### Author Declarations

The University of Illinois at Chicago Institutional Review Board deemed the use of the database for this study exempt.

